# Unequal Burden and Unequal Access: Socioeconomic Disparities in Tuberculosis Diagnosis and Out-of-Pocket Expenditures in South Africa and Lesotho

**DOI:** 10.64898/2025.12.04.25341662

**Authors:** Harsh Vivek Harkare, Anna Verjans, Shriya Misra, Mashaete Kamele, Thandanani Madonsela, Alastair van Heerden, Marina Antillon, Bart K.M. Jacobs, Lutgarde Lynen, Curdin Brugger, Johanna Kurscheid, Tracy R. Glass, Fiona Vanobberghen, Shannon Bosman, Irene Ayakaka, Aita Signorell, Klaus Reither, Fabrizio Tediosi

## Abstract

Tuberculosis (TB) remains a major public health challenge in Southern Africa, where socioeconomic disparities continue to influence access to care, diagnostic pathways, and treatment outcomes. This study examines how wealth, comorbidities, and household economic vulnerability drive TB diagnosis and access to care in South Africa and Lesotho, with the aim of informing more equitable TB interventions. We analysed data from a community-based diagnostic trial comparing two TB screening strategies and assessed the socioeconomic and clinical profiles of individuals diagnosed via active case finding (ACF). Additional participants were enrolled via a passive case finding approach to allow for comparison. We examined associations between socioeconomic status (SES) and both TB-related outcomes (diagnosis pathway, treatment status) and non-TB economic outcomes (TB-associated out-of-pocket expenditures (OOP) and income change) among 417 participants, applying logistic regression and stratified analyses Higher SES was associated with linkage to care among TB-positive individuals, as indicated by the presence of a documented TB record. Participants from higher SES groups incurred greater absolute OOP expenditures, particularly in South Africa. In Lesotho, income loss and borrowing were uncommon but disproportionately affected those in the lowest SES group, who also experienced a higher financial burden relative to income. Higher educational attainment and HIV-positive status were associated with greater odds of being diagnosed with TB through passive case finding and of receiving TB treatment. No significant changes in household income were observed between baseline and follow-up 56 ±6 days later among ACF-identified participants. Overall, only 3.3% of those reporting OOP costs experienced catastrophic health expenditure (>20% of monthly income).

**Funding:** This project is part of the EDCTP2 programme (grant number RIA2018D-2498; TB TRIAGE+) supported by the European Union.

**Trial registration:** ClinicalTrials.gov (NCT05526885), South African National Clinical Trials Register (SANCTR; DOH-27-092022-8096).

## Introduction

Tuberculosis (TB) remains a leading cause of morbidity and mortality in Southern Africa, with South Africa and Lesotho bearing some of the highest burdens globally [1–4]. While clinical and diagnostic advancements have improved TB control efforts, socioeconomic disparities continue to shape access to care, diagnosis pathways, and treatment outcomes [5]. A growing body of evidence underscores the role of social determinants in driving TB incidence, both through direct and indirect pathways [6]. In particular, out-of-pocket payments (OOP), household income shocks, and the interplay between TB and non-communicable diseases (NCDs) may deepen vulnerability to treatment delays, disease progression, and worsening poverty among affected populations - especially in resource-constrained settings. Reducing poverty due to illness and mitigating catastrophic health expenditure is a key priority under the Sustainable Development Goals (SDGs), specifically SDG 1 (No Poverty) and SDG 3 (Good Health and Well-being) [7]. Understanding how these factors vary across wealth strata is critical to designing equitable health interventions.

This study investigates the socioeconomic and clinical profiles of individuals diagnosed with TB in KwaZulu-Natal, South Africa and Butha Buthe district, Lesotho, exploring how socioeconomic characteristics influence diagnostic pathways, co-morbidities, and the economic burden of care. By examining household income changes, OOP expenditures, and diagnostic pathway (active case finding (ACF) vs passive case finding (PCF) approaches), we aim to identify patterns of vulnerability that can inform more targeted, context-sensitive approaches to TB care and social protection.

## Materials and Methods

### Study design and setting

This study is nested within a large, pragmatic, community-based diagnostic trial conducted in Butha Buthe district in Lesotho and KwaZulu-Natal in South Africa, designed to evaluate the effectiveness and cost-effectiveness of two TB screening strategies - Computer-Aided Detection for TB (CAD4TB) alone versus CAD4TB combined with C-reactive protein assay (CRP) - in detecting tuberculosis in high-burden, underserved communities. Villages in communities with high TB burden and limited access to healthcare and TB screening were randomly selected from an eligible sampling frame, and all eligible adults residing in these villages were invited to participate. The systematic identification of TB cases through outreach screening activities is called active case finding (ACF), as opposed to passive case finding (PCF), where individuals are diagnosed after presenting with symptoms at a health facility. Data on ACF participants were obtained directly from the trial, whereas a separate subgroup of individuals diagnosed through PCF was recruited at health facilities to allow comparison between the two groups. The trial not only seeks to improve case detection through innovative, decentralised screening but also aims to examine the broader social and economic implications of a TB diagnosis at the household level. Recognising that TB disproportionately affects individuals in socioeconomically vulnerable settings, the study incorporates comprehensive assessments of household wealth, OOP expenditures, and income changes following diagnosis. Further information on our screening strategies and data collection is presented in the supplement. Details of the study setting, eligibility criteria, and the overall trial are outlined in the protocol [8].

### Data

Data were collected over a two-year period, beginning on September 15, 2022, in Lesotho and on October 1, 2022, in South Africa, and concluding on August 31, 2024, across both sites. All participants provided written informed consent prior to enrolment. For illiterate participants, consent was obtained via fingerprint in the presence of an impartial witness. Consent forms were translated into Sesotho and Zulu, and interviews were conducted in private settings. No minors were enrolled in the study.

The study questionnaire was administered twice: Once at baseline, defined as the day of screening or the following day; and then again at follow-up, conducted 56 ± 6 days after baseline. To assess the financial burden associated with TB, structured interviews were conducted by trained field workers using REDCap electronic data capture software [9]. Participants were categorised into four distinct groups, stratified by case-finding approach and TB status:

1. ACF TB-positive: Participants testing positive for TB by Xpert Ultra and/or TB LAM (including trace results) during ACF.
2. ACF suspected TB: Participants who were clinically suspected of TB at baseline or unable to produce sputum but were on TB treatment at follow-up (based on self-report or TB records identified through ACF).
3. PCF TB-positive: A sample of adults (n = 150) newly diagnosed with TB within the past month through PCF at health facilities in the study area.
4. ACF TB-negative: A randomly selected subset of individuals from ACF-identified TB-negative households (n = 160).

OOP and household economic data were collected only from participants belonging to one of these four predefined groups, who together constituted the full eligible population for the socioeconomic sub-study. For the first three groups, we collected information on OOP expenditures and indirect costs incurred due to TB symptoms up to the point of diagnosis. These included costs for medical consultations, transportation, food, lost workdays, and any prior hospitalisations related to TB. In addition, for the first two groups, additional data were collected on (i) OOP expenditures related to TB diagnosis and treatment, (ii) coping strategies used to manage TB-related costs, and (iii) expenditures for outpatient medical visits during follow-up. Across all four groups, we collected baseline data on monthly household income and household assets to assess household wealth. These data enabled comparisons between TB-positive and TB-negative participants within ACF, and between ACF- and PCF-identified cases.

### Analysis

We derived a household wealth index for the study population using the International Wealth Index (IWI) methodology, which is a harmonised, asset-based measure of socioeconomic status validated for use across low- and middle-income countries [10]. The index was constructed using the principal component analysis (PCA) method applied to a standardised set of variables capturing asset ownership, housing quality, and access to basic services [11]. To account for contextual differences in asset ownership and infrastructure, the index was constructed separately for South Africa and Lesotho. Participants were classified into three groups of socioeconomic status (SES) - low, middle, and high - based on national IWI cut-offs provided by the Global Data Lab (GDL) and aligned with standard conventions [12, 13]. This approach improves comparability with population-representative distributions, given that our study sample was purposively designed to reach underserved populations and may not be nationally representative. Participants who did not respond to, or were unable to complete, the health economic questionnaire required to construct the household wealth index were excluded from the analysis. Further details are provided in the supplement.

We used logistic regression models to examine whether socioeconomic status was associated with TB-related outcomes, including having a TB record at a health facility, being diagnosed via ACF, receiving treatment, or being identified through PCF. The presence of a TB record is identified at follow-up and reflects linkage to a healthcare facility following diagnosis via ACF and offers complementary information on engagement with care beyond initial TB identification. We also assessed the association of SES with self-reported co-morbid conditions, specifically hypertension, diabetes, and heart disease. Each logistic regression model was adjusted for age, sex, HIV status, education level, and previous TB history. These were selected a priori based on published evidence regarding socioeconomic and clinical determinants of TB risk, access to care, and co-morbidities in Southern Africa, allowing a clearer assessment of whether socioeconomic status is independently associated with each outcome [2]. In addition, we also test where SES was jointly associated with each outcome using Wald tests to evaluate the overall effect of SES across all categories. Age was assessed for non-linearity using spline terms and finally modelled as a continuous linear variable. We scaled age in 5-year increments, with coefficients reflecting the change in odds per 5-year increase. Model results are presented as adjusted odds ratios (ORs) with corresponding 95% confidence intervals (CI).

To explore the economic burden associated with TB, we conducted multiple analyses. We compared overall OOP expenditures for TB-related care among those who incurred any OOP payments - including food, travel, consultations, laboratory tests, medicines, radiography, hospitalisation, and procedures - across socioeconomic groups using the Kruskal-Wallis test. Pairwise comparisons of the socioeconomic groups were performed using Wilcoxon rank-sum tests with Bonferroni corrections. We also examined differences in OOP spending by diagnostic pathway and by country, and evaluated supplementary TB-specific expenditures such as nutritional supplements that participants would not have purchased in the absence of TB. The relative financial burden of TB-related care was calculated as the proportion of monthly household income spent on OOP costs at baseline. We assessed whether participants incurred catastrophic healthcare expenditure (CHE), defined as spending more than 20% of monthly income on OOP. Differences in relative burden across SES was assessed using the Kruskal-Wallis and Wilcoxon rank-sum tests, as appropriate.

We evaluated changes in household income between baseline and follow-up (Day 56 ± 6 days) using the Wilcoxon signed-rank test. Differences in income between baseline and follow-up were also assessed by sex, socioeconomic group, TB diagnosis status, and perceived financial impact of TB. Income comparisons across categorical groups were conducted using Wilcoxon rank-sum or Kruskal-Wallis tests, depending on the number of comparison groups. Finally, we explored coping strategies for financing OOP expenditures as reported by participants, including borrowing money and selling assets, and summarised these descriptively.

All income and expenditure values were converted to 2023 USD from ZAR/LSL using the exchange rate at the time of data collection (1 ZAR = 0.05424 USD) [14]. Analyses were performed in Stata (version 18.5) and R (version 4.4.1).

### Ethics Statement

The study protocol was approved by the National Health Research Ethics Committee in Lesotho (NH-REC, ID52-2022), the Human Sciences Research Council Research Ethics Committee in South Africa (HSRC REC, REC 2/23/09/20), the Provincial Health Research Committee of the Department of Health of KwaZulu-Natal (KZ_202209_022), and the Swiss Ethics Committee Northwest and Central Switzerland (EKNZ, AO_2022–00044). Written informed consent was obtained from all study participants prior to enrolment [8].

## Results

### Study participants and characteristics

Figure 1 illustrates the flow of participants across the four groups. A total of 422 trial participants eligible for OOP data collection, of whom 417 had complete asset ownership information and were included in the analytic sample - 199 from South Africa and 218 from Lesotho. Among these, 112 participants were Xpert Ultra-positive (Group 1), including 35 with trace results, while no participants tested positive on TB LAM in our analytical subset. None of the participants in our analytical subsample met the criteria for suspected TB (Group 2). After excluding five individuals due to missing asset ownership data, 109 participants were included from the ACF TB-positive group (Group 1), 149 participants were included from the PCF group (Group 3), and 159 from the TB-negative group (Group 4).

**Figure 1.**
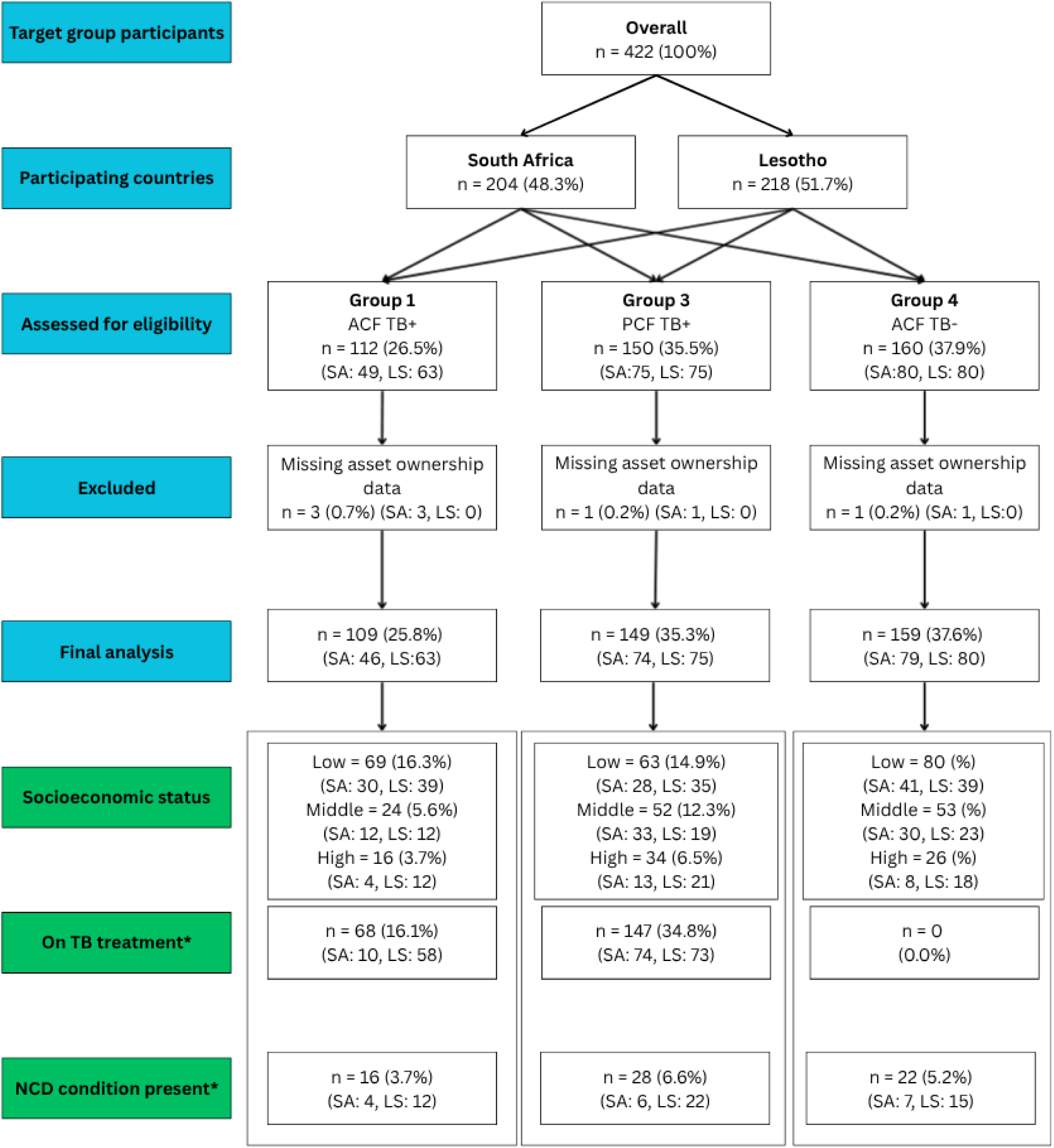
**CONSORT flowchart of participants included in the analysis** *Note: On TB treatment refers to TB treatment status at follow-up as per a documented TB record. NCD condition present refers to either hypertension, diabetes, or heart disease.

Table 1 presents the socioeconomic and clinical characteristics of study participants in South Africa (n = 199) and Lesotho (n = 218), stratified by SES group. In South Africa, the median age was 37.5 years (IQR: 27-48), and the prevalence of self-reported NCDs (includes hypertension, diabetes, and heart disease only) was 8.5%, limited to the low and middle SES groups. Among TB-positive participants in the low SES group, a greater proportion were diagnosed via ACF (30.3%) compared to PCF (28.2%), whereas in the high SES group, diagnosis via PCF was more common (52.0%) than ACF (16.0%). A documented TB record was found for 34.7% of ACF-identified TB-positive participants (n = 16), suggesting that few people engaged with a healthcare facility for diagnosis confirmation or treatment initiation. However, a substantial number of participants experienced missed referrals during the trial, which has direct implications on having a documented TB record. Among these ACF-identified TB-positive individuals, 62.5% (n = 10) were reported to be on TB treatment. In Lesotho, the average IWI score ranged from 20.5 in the low group to 59.96 in the high SES group. The median age was 55 years (IQR: 38-69) and NCDs were reported by 22.4% of participants, highest in the low (25.6%) and high (27.4%) SES? groups. Among TB-positive individuals (n = 138), 96.8% had a documented TB record, and treatment coverage remained high overall at 95.0%, with limited variation across socioeconomic groups.

**Table 1.**
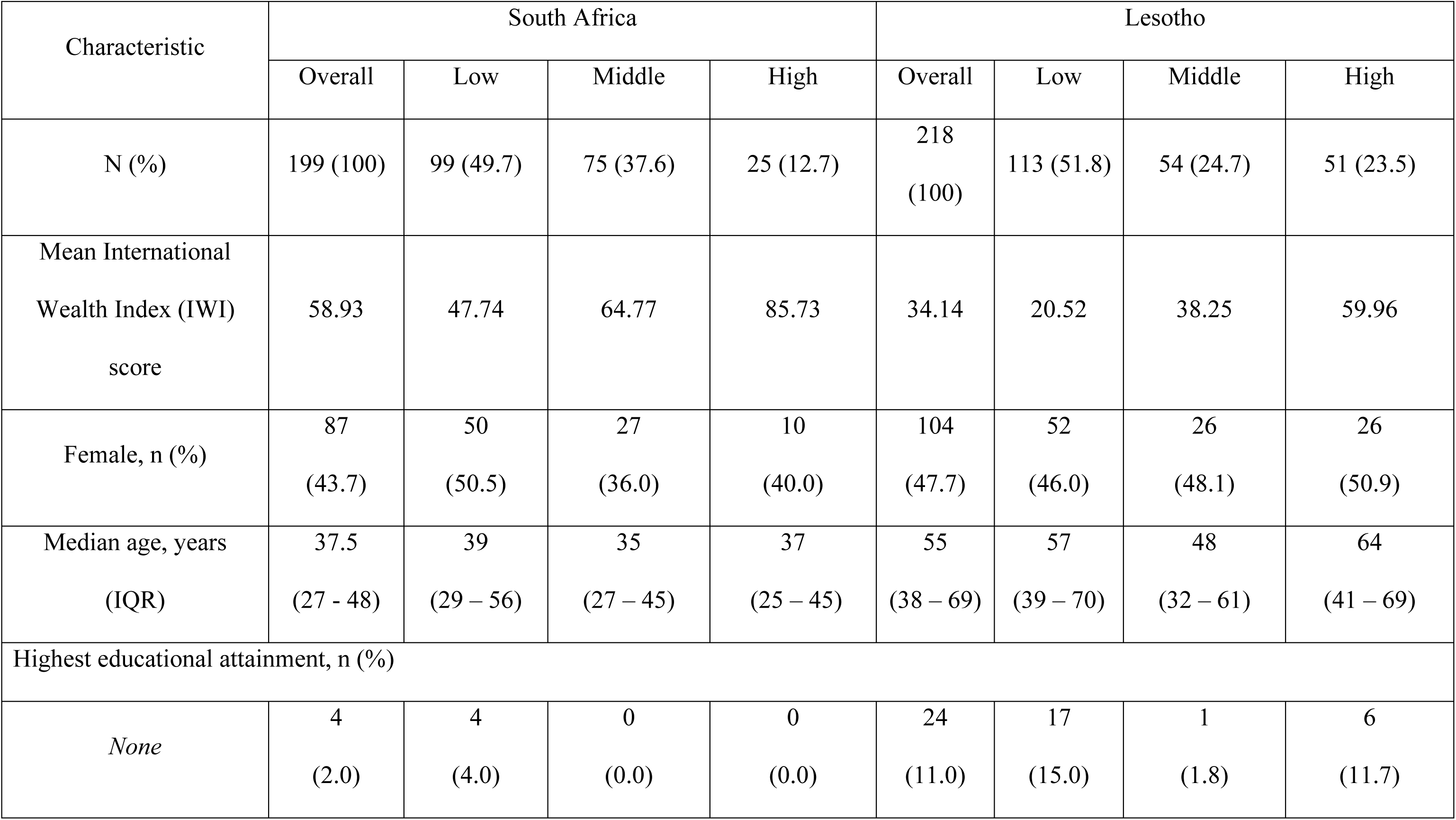
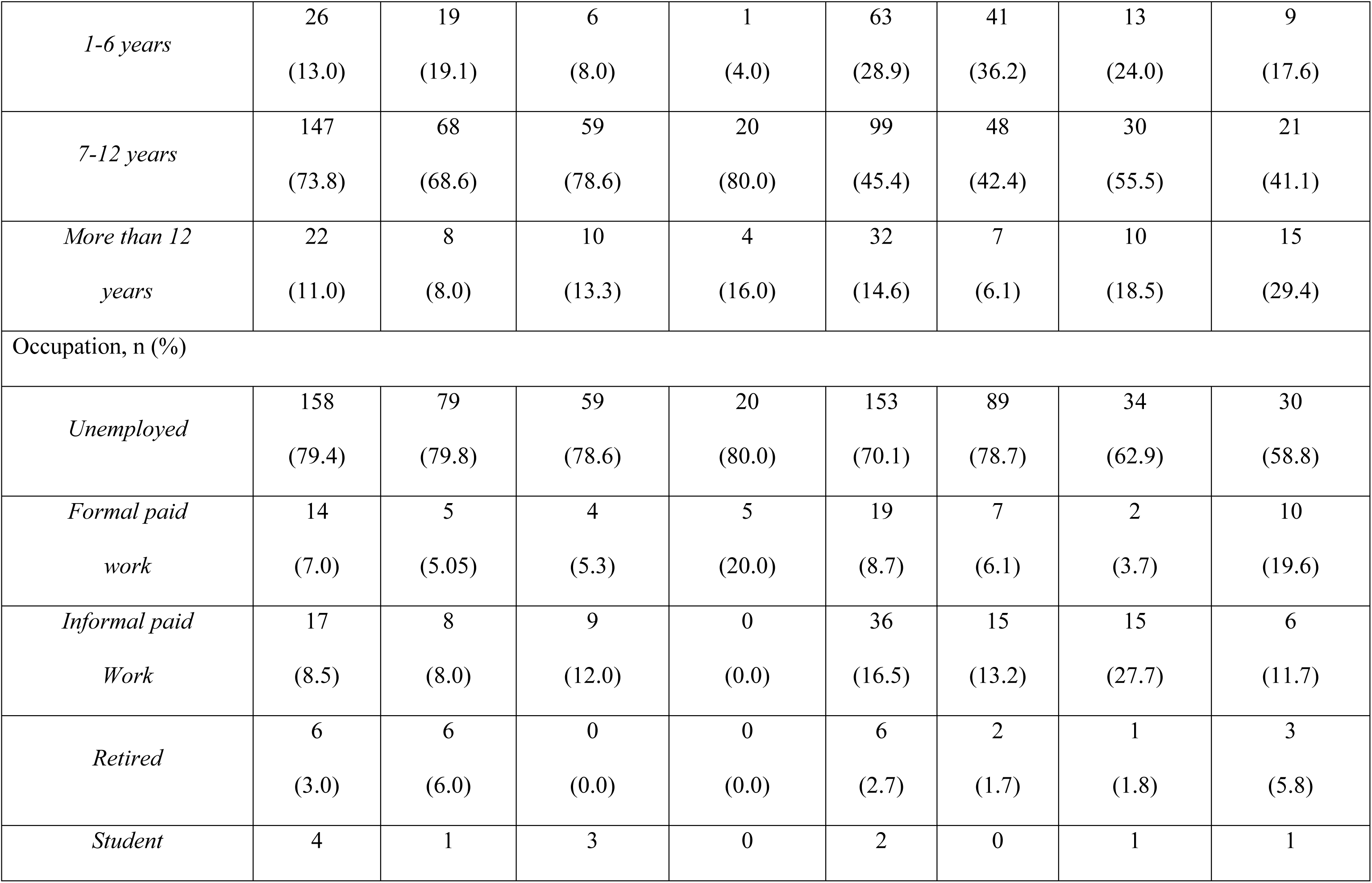
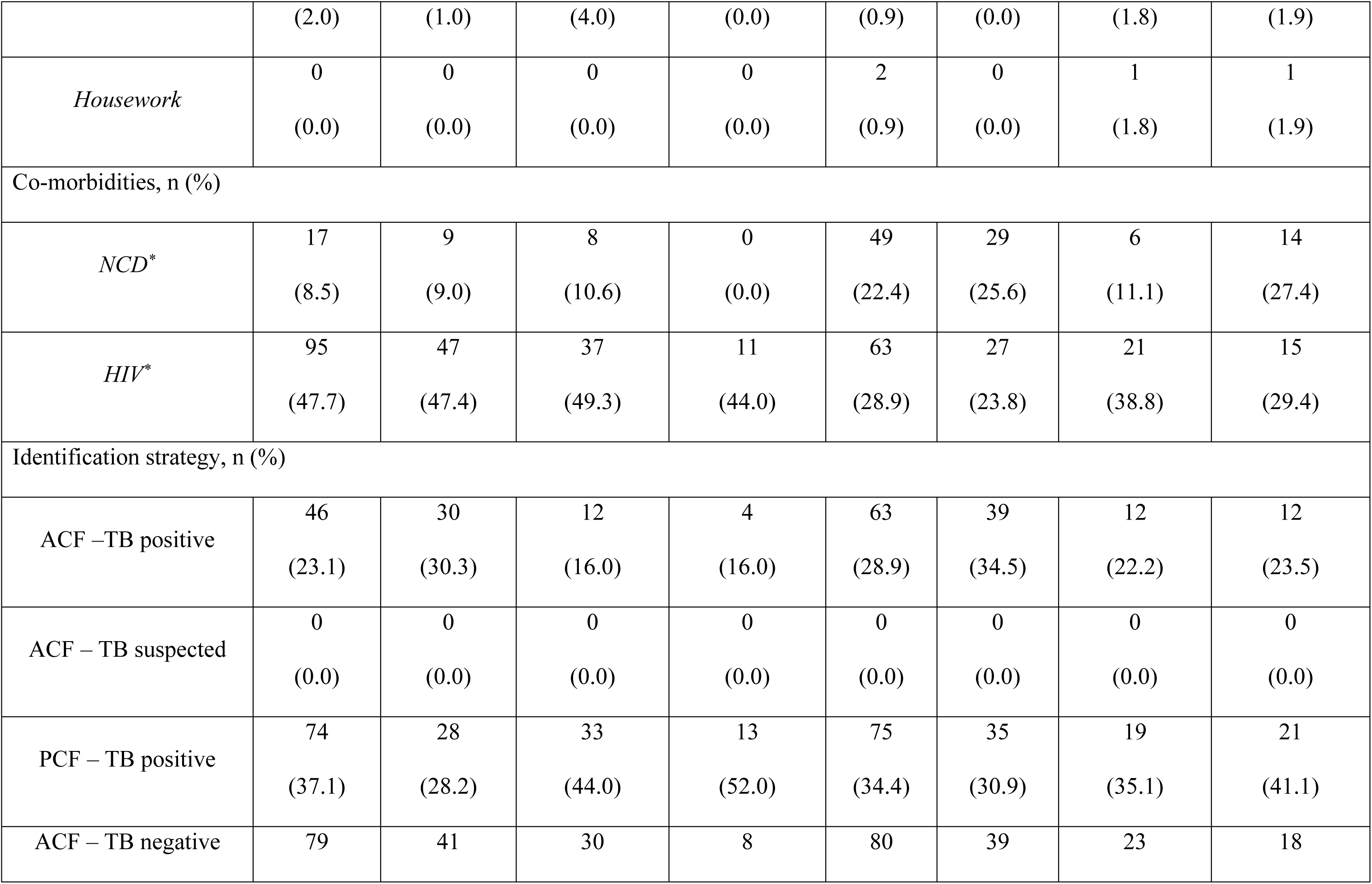
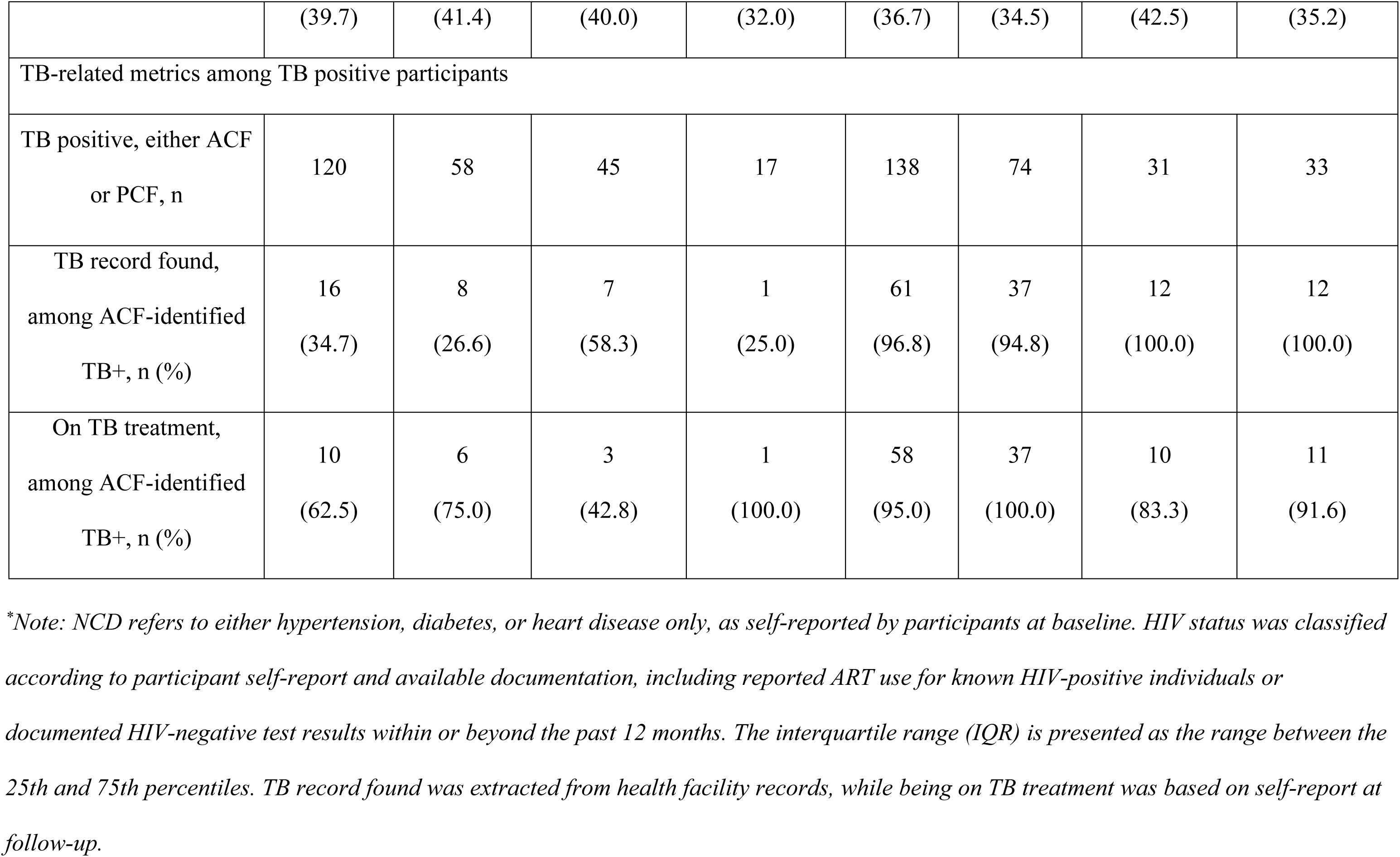
Socioeconomic and clinical characteristics of participants by country and socioeconomic group.

| Characteristic | South Africa |  |  |  | Lesotho |  |  |  |
| --- | --- | --- | --- | --- | --- | --- | --- | --- |
|  | Overall | Low | Middle | High | Overall | Low | Middle | High |
| N (%) | 199 (100) | 99 (49.7) | 75 (37.6) | 25 (12.7) | 218<br>(100) | 113 (51.8) | 54 (24.7) | 51 (23.5) |
| Mean International<br>Wealth Index (IWI)<br>score | 58.93 | 47.74 | 64.77 | 85.73 | 34.14 | 20.52 | 38.25 | 59.96 |
| Female, n (%) | 87<br>(43.7) | 50<br>(50.5) | 27<br>(36.0) | 10<br>(40.0) | 104<br>(47.7) | 52<br>(46.0) | 26<br>(48.1) | 26<br>(50.9) |
| Median age, years<br>(IQR) | 37.5<br>(27 - 48) | 39<br>(29 – 56) | 35<br>(27 – 45) | 37<br>(25 – 45) | 55<br>(38 – 69) | 57<br>(39 – 70) | 48<br>(32 – 61) | 64<br>(41 – 69) |
| Highest educational attainment, n (%) |  |  |  |  |  |  |  |  |
| *None* | 4<br>(2.0) | 4<br>(4.0) | 0<br>(0.0) | 0<br>(0.0) | 24<br>(11.0) | 17<br>(15.0) | 1<br>(1.8) | 6<br>(11.7) |
| *1-6 years* | 26<br>(13.0) | 19<br>(19.1) | 6<br>(8.0) | 1<br>(4.0) | 63<br>(28.9) | 41<br>(36.2) | 13<br>(24.0) | 9<br>(17.6) |
| *7-12 years* | 147<br>(73.8) | 68<br>(68.6) | 59<br>(78.6) | 20<br>(80.0) | 99<br>(45.4) | 48<br>(42.4) | 30<br>(55.5) | 21<br>(41.1) |
| *More than 12*<br>*years* | 22<br>(11.0) | 8<br>(8.0) | 10<br>(13.3) | 4<br>(16.0) | 32<br>(14.6) | 7<br>(6.1) | 10<br>(18.5) | 15<br>(29.4) |
| Occupation, n (%) |  |  |  |  |  |  |  |  |
| *Unemployed* | 158<br>(79.4) | 79<br>(79.8) | 59<br>(78.6) | 20<br>(80.0) | 153<br>(70.1) | 89<br>(78.7) | 34<br>(62.9) | 30<br>(58.8) |
| *Formal paid*<br>*work* | 14<br>(7.0) | 5<br>(5.05) | 4<br>(5.3) | 5<br>(20.0) | 19<br>(8.7) | 7<br>(6.1) | 2<br>(3.7) | 10<br>(19.6) |
| *Informal paid*<br>*Work* | 17<br>(8.5) | 8<br>(8.0) | 9<br>(12.0) | 0<br>(0.0) | 36<br>(16.5) | 15<br>(13.2) | 15<br>(27.7) | 6<br>(11.7) |
| *Retired* | 6<br>(3.0) | 6<br>(6.0) | 0<br>(0.0) | 0<br>(0.0) | 6<br>(2.7) | 2<br>(1.7) | 1<br>(1.8) | 3<br>(5.8) |
| *Student* | 4 | 1 | 3 | 0 | 2 | 0 | 1 | 1 |
|  | (2.0) | (1.0) | (4.0) | (0.0) | (0.9) | (0.0) | (1.8) | (1.9) |
| *Housework* | 0<br>(0.0) | 0<br>(0.0) | 0<br>(0.0) | 0<br>(0.0) | 2<br>(0.9) | 0<br>(0.0) | 1<br>(1.8) | 1<br>(1.9) |
| Co-morbidities, n (%) |  |  |  |  |  |  |  |  |
| *NCD\** | 17<br>(8.5) | 9<br>(9.0) | 8<br>(10.6) | 0<br>(0.0) | 49<br>(22.4) | 29<br>(25.6) | 6<br>(11.1) | 14<br>(27.4) |
| *HIV\** | 95<br>(47.7) | 47<br>(47.4) | 37<br>(49.3) | 11<br>(44.0) | 63<br>(28.9) | 27<br>(23.8) | 21<br>(38.8) | 15<br>(29.4) |
| Identification strategy, n (%) |  |  |  |  |  |  |  |  |
| ACF –TB positive | 46<br>(23.1) | 30<br>(30.3) | 12<br>(16.0) | 4<br>(16.0) | 63<br>(28.9) | 39<br>(34.5) | 12<br>(22.2) | 12<br>(23.5) |
| ACF – TB suspected | 0<br>(0.0) | 0<br>(0.0) | 0<br>(0.0) | 0<br>(0.0) | 0<br>(0.0) | 0<br>(0.0) | 0<br>(0.0) | 0<br>(0.0) |
| PCF – TB positive | 74<br>(37.1) | 28<br>(28.2) | 33<br>(44.0) | 13<br>(52.0) | 75<br>(34.4) | 35<br>(30.9) | 19<br>(35.1) | 21<br>(41.1) |
| ACF – TB negative | 79 | 41 | 30 | 8 | 80 | 39 | 23 | 18 |
|  | (39.7) | (41.4) | (40.0) | (32.0) | (36.7) | (34.5) | (42.5) | (35.2) |
| TB-related metrics among TB positive participants |  |  |  |  |  |  |  |  |
| TB positive, either ACF<br>or PCF, n | 120 | 58 | 45 | 17 | 138 | 74 | 31 | 33 |
| TB record found,<br>among ACF-identified<br>TB+, n (%) | 16<br>(34.7) | 8<br>(26.6) | 7<br>(58.3) | 1<br>(25.0) | 61<br>(96.8) | 37<br>(94.8) | 12<br>(100.0) | 12<br>(100.0) |
| On TB treatment,<br>among ACF-identified<br>TB+, n (%) | 10<br>(62.5) | 6<br>(75.0) | 3<br>(42.8) | 1<br>(100.0) | 58<br>(95.0) | 37<br>(100.0) | 10<br>(83.3) | 11<br>(91.6) |

*\*Note: NCD refers to either hypertension, diabetes, or heart disease only, as self-reported by participants at baseline. HIV status was classified* *according to participant self-report and available documentation, including reported ART use for known HIV-positive individuals or* *documented HIV-negative test results within or beyond the past 12 months. The interquartile range (IQR) is presented as the range between the* *25th and 75th percentiles. TB record found was extracted from health facility records, while being on TB treatment was based on self-report at* *follow-up.*

The socioeconomic characteristics of study participants by TB status, by diagnostic pathway and participant groups, and by reporting of OOP expenditure at baseline, are provided in Tables S1-3.

### TB- and NCD-related outcomes across SES groups

Table 2 presents results from five logistic regression models evaluating key TB and NCD-related outcomes by sociodemographic and clinical characteristics. Among the study population, 77 participants had a documented TB record at a health facility (indicating linkage to care), 109 were diagnosed through ACF, 215 had documented evidence of TB treatment initiation, and 66 were identified as having an NCD.

**Table 2.**
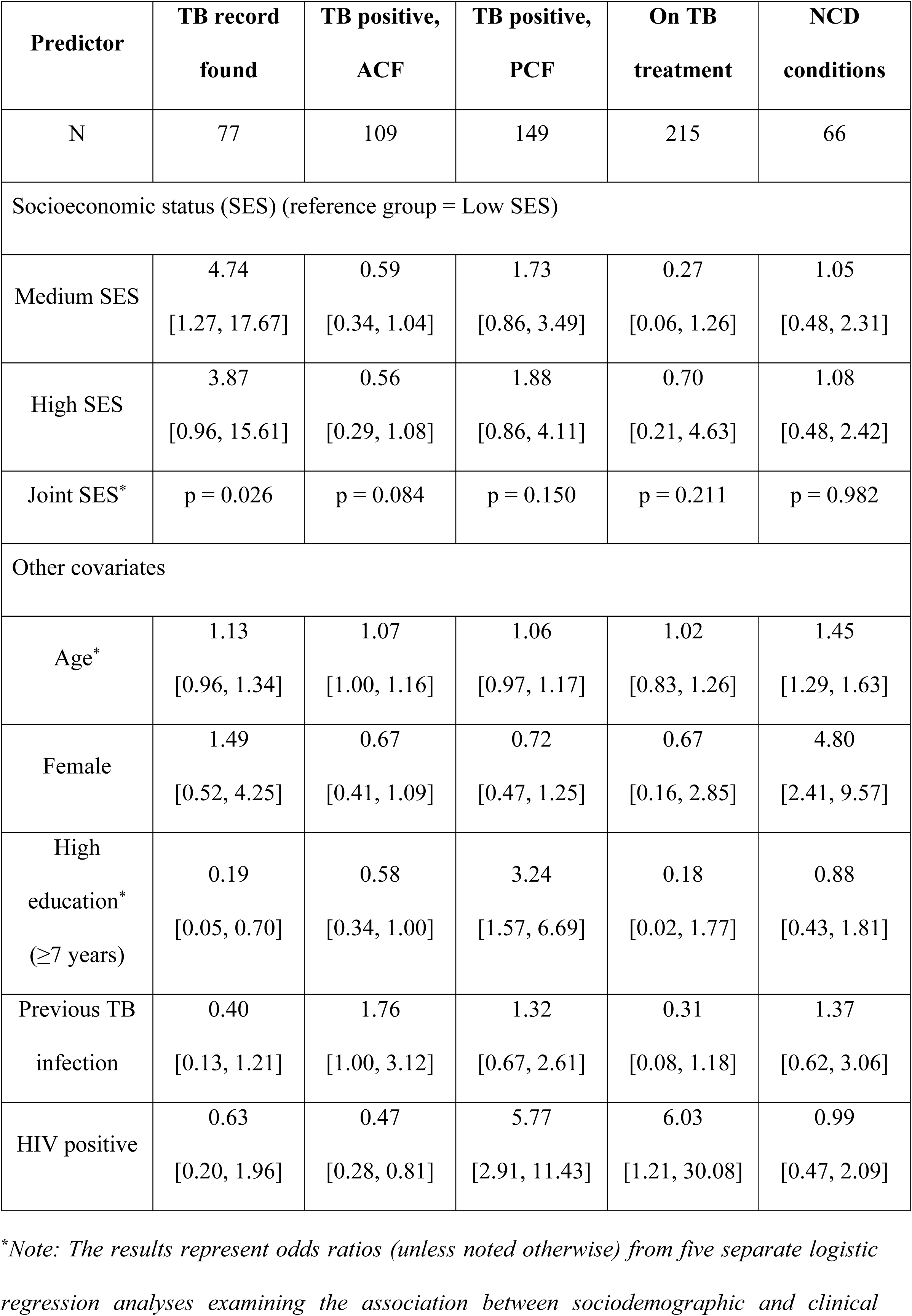
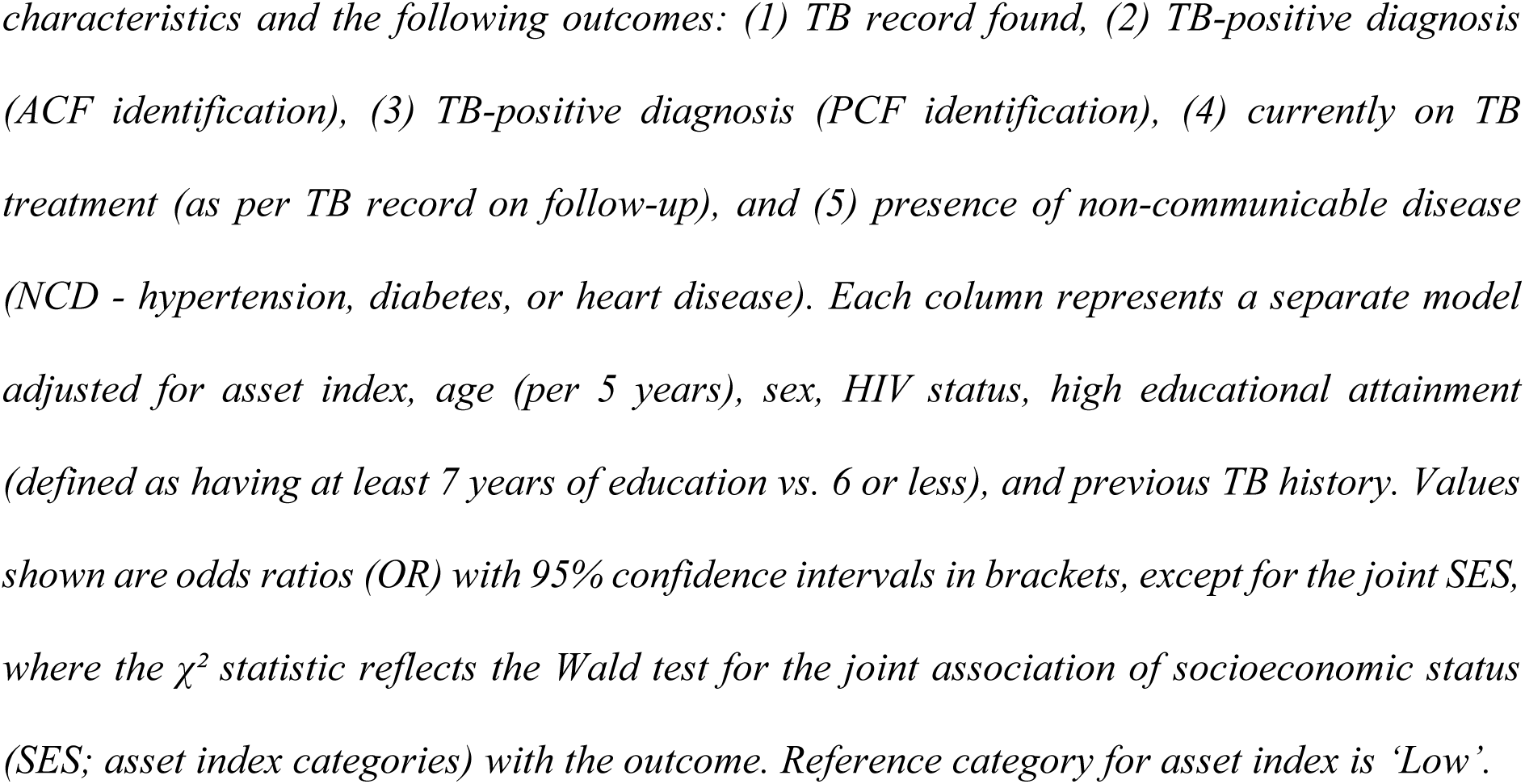
Logistic regression results: adjusted odds ratios and joint tests for SES.

| **Predictor** | **TB record found** | **TB positive, ACF** | **TB positive, PCF** | **On TB treatment** | **NCD conditions** |
| --- | --- | --- | --- | --- | --- |
| N | 77 | 109 | 149 | 215 | 66 |
| Socioeconomic status (SES) (reference group = Low SES) |  |  |  |  |  |
| Medium SES | 4.74<br>[1.27, 17.67] | 0.59<br>[0.34, 1.04] | 1.73<br>[0.86, 3.49] | 0.27<br>[0.06, 1.26] | 1.05<br>[0.48, 2.31] |
| High SES | 3.87<br>[0.96, 15.61] | 0.56<br>[0.29, 1.08] | 1.88<br>[0.86, 4.11] | 0.70<br>[0.21, 4.63] | 1.08<br>[0.48, 2.42] |
| Joint SES* | p = 0.026 | p = 0.084 | p = 0.150 | p = 0.211 | p = 0.982 |
| Other covariates |  |  |  |  |  |
| Age* | 1.13<br>[0.96, 1.34] | 1.07<br>[1.00, 1.16] | 1.06<br>[0.97, 1.17] | 1.02<br>[0.83, 1.26] | 1.45<br>[1.29, 1.63] |
| Female | 1.49<br>[0.52, 4.25] | 0.67<br>[0.41, 1.09] | 0.72<br>[0.47, 1.25] | 0.67<br>[0.16, 2.85] | 4.80<br>[2.41, 9.57] |
| High education*<br>(≥7 years) | 0.19<br>[0.05, 0.70] | 0.58<br>[0.34, 1.00] | 3.24<br>[1.57, 6.69] | 0.18<br>[0.02, 1.77] | 0.88<br>[0.43, 1.81] |
| Previous TB infection | 0.40<br>[0.13, 1.21] | 1.76<br>[1.00, 3.12] | 1.32<br>[0.67, 2.61] | 0.31<br>[0.08, 1.18] | 1.37<br>[0.62, 3.06] |
| HIV positive | 0.63<br>[0.20, 1.96] | 0.47<br>[0.28, 0.81] | 5.77<br>[2.91, 11.43] | 6.03<br>[1.21, 30.08] | 0.99<br>[0.47, 2.09] |

\*Note: The results represent odds ratios (unless noted otherwise) from five separate logistic regression analyses examining the association between sociodemographic and clinical
characteristics and the following outcomes: (1) TB record found, (2) TB-positive diagnosis (ACF identification), (3) TB-positive diagnosis (PCF identification), (4) currently on TB treatment (as per TB record on follow-up), and (5) presence of non-communicable disease (NCD - hypertension, diabetes, or heart disease). Each column represents a separate model adjusted for asset index, age (per 5 years), sex, HIV status, high educational attainment (defined as having at least 7 years of education vs. 6 or less), and previous TB history. Values shown are odds ratios (OR) with 95% confidence intervals in brackets, except for the joint SES, where the $\chi^2$ statistic reflects the Wald test for the joint association of socioeconomic status (SES; asset index categories) with the outcome. Reference category for asset index is 'Low'.

Higher SES was associated with having a TB record in the health facility, reflecting stronger linkage to care following diagnosis. Compared to the low SES group, individuals in the middle and high SES groups had higher odds of having a documented TB record (OR = 4.74, 95% CI: 1.27-17.67 and OR = 3.87, 95% CI: 0.96-15.61, respectively). The overall association between SES and having a TB record was further supported by a joint Wald test (χ²(2) = 7.31, p = 0.026). No joint association was observed between SES and ACF or PCF diagnosis, TB treatment initiation, or NCD diagnosis.

Older age and female sex were associated with higher odds of having an NCD condition (OR = 1.45, 95% CI: 1.29, 1.63; p < 0.001, and OR = 4.80, 95% CI: 2.41, 9.57; p < 0.001respectively). Having 7 years of education or more was associated with lower odds of having a TB record (OR = 0.19, 95% CI: 0.05, 0.70; p = 0.012), lower odds of TB positivity via ACF (OR = 0.58, 95% CI: 0.34, 1.00; p = 0.059) and higher odds of TB positivity diagnosed via PCF (OR = 3.24, 95% CI: 1.57, 6.69; p = 0.001). HIV-positive participants had lower odds of being TB-positive via ACF (OR = 0.47, 95% CI: 0.28, 0.81; p = 0.007), but higher odds of being diagnosed through PCF (OR = 5.77, 95% CI: 2.91, 11.43; p < 0.001) and being on TB treatment at follow-up (OR = 6.03, 95% CI: 1.21, 30.08; p = 0.028).

In country-specific analyses (Tables S4-5), PCF-based TB diagnosis was more common among individuals in the high socioeconomic group in South Africa, with an overall association between SES and PCF diagnosis (p = 0.044). Those in the middle group were less likely to be diagnosed with TB via ACF.

### Impact on OOP expenditure by SES group

#### To receive TB diagnosis at baseline

Of the 258 participants who tested positive for TB, 34.4% (n = 89) reported having sought care prior to diagnosis and completed the OOP expenditure form. These included 59 individuals from South Africa (25 low SES, 24 middle, 10 high) and 30 from Lesotho (15 low, 4 middle, 21 high). Within each country, those who reported OOP data were broadly similar to non-reporting participants in their socioeconomic and clinical characteristics with some differences (Table S3).

Figure 2 shows average OOP expenditure from the most recent TB-related healthcare visit, reported by TB-positive participants who sought care prior to diagnosis at baseline. Results are stratified by socioeconomic group and country and disaggregated by expenditure category. OOP expenditure stratified by diagnostic pathway and country are presented in Figure S1.

**Figure 2.**
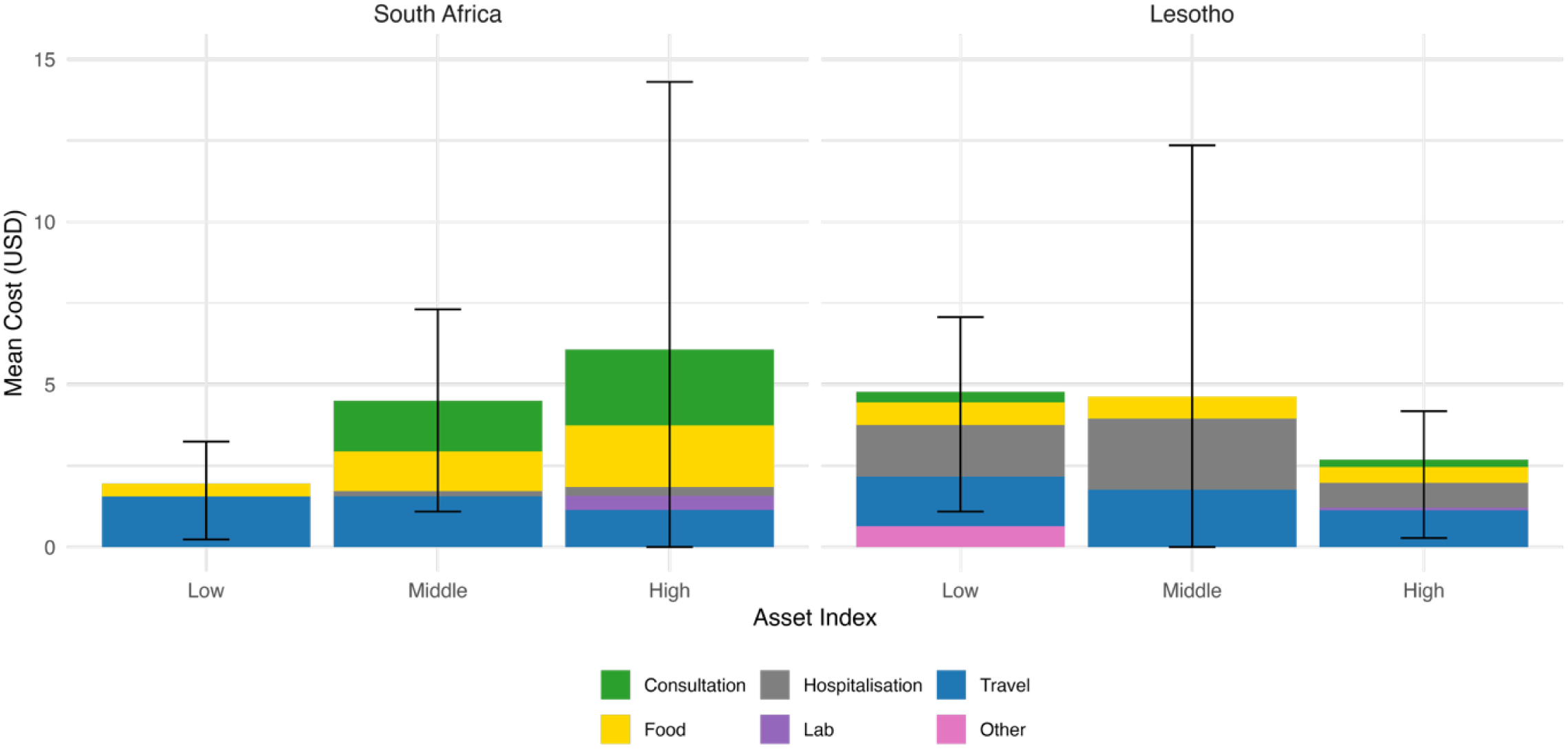
**Average out-of-pocket expenditure to seek TB diagnosis at baseline by socioeconomic group, country, and expenditure category (ACF or PCF).** Note: Among TB-positive participants identified via ACF or PCF (n = 258), those who reported seeking care prior to diagnosis (n = 89) were asked about out-of-pocket costs from their most recent TB-related facility visit. The stacked bars illustrate the composition of average expenditure across six cost categories. Black vertical lines represent 95% confidence intervals around the total mean cost per group. Costs are collected in ZAR/LSL and converted to USD.

In South Africa, mean OOP expenditure was highest in the high SES group ($6.05), followed by the middle ($4.50) and low SES groups ($2.00). In Lesotho, costs were highest in the low SES group ($4.80), with lower means in the middle ($4.60) and high ($2.60) SES groups. No participants reported expenditures for radiography, imaging, or medications. However, confidence intervals were wide in certain groups, particularly among high SES participants in South Africa and lower-income participants in Lesotho. This indicates substantial variability in reported OOP expenditures.

In South Africa, spending was largest among individuals in the high SES group, driven by consultation and travel costs. By contrast, OOP spending in Lesotho was highest among the poorest group, with hospitalisation, travel, and other expenses such as nutritional supplements and medical procedures contributing most to the total. Most individuals in South Africa sought primary care at a primary health centre (81.3%), whereas in Lesotho, the majority sought primary care at a hospital (58.0%). All visits to private healthcare facilities were reported by participants in the middle and high SES group in South Africa (n = 4), and none in Lesotho. Table S6 shows average OOP among TB positive participants reporting and incurring costs (i.e. OOP > 0). In South Africa, costs increased with SES, whereas no clear trend was observed in Lesotho. Mean costs were higher among PCF-diagnosed individuals in Lesotho but not in South Africa. The observed differences were not statistically significant (Table S7).

### To access TB treatment at follow-up

Figure 3 presents the mean OOP expenditure on TB-related follow-up costs incurred by TB-positive participants identified via ACF after diagnosis as reported at day 56, stratified by country and SES group. Overall, data were available for 65 participants, including 15 from South Africa and 50 from Lesotho. Across both countries, food and travel consistently represented the largest share of total follow-up expenditure. In South Africa, mean OOP costs were highest in the high SES group ($5.80), followed by the low ($4.50) and middle ($2.30) groups, primarily driven by spending on travel, food, and consultations. In Lesotho, mean expenditures were lower and relatively consistent across SES groups, ranging from $1.90 in the low SES group to $2.50 in the high SES group, with food and travel comprising the bulk of expenses. However, substantial variation in OOP spending was observed as denoted by the wide confidence intervals, especially in South Africa. Overall, 5 participants (6.0%) reported purchasing nutritional supplements such as vitamins outside of their regular diet due to their TB illness at follow-up (2 in SA, 3 in LS), with a median expenditure of $13.56 (IQR: $5.42 - 17.62).

**Figure 3.**
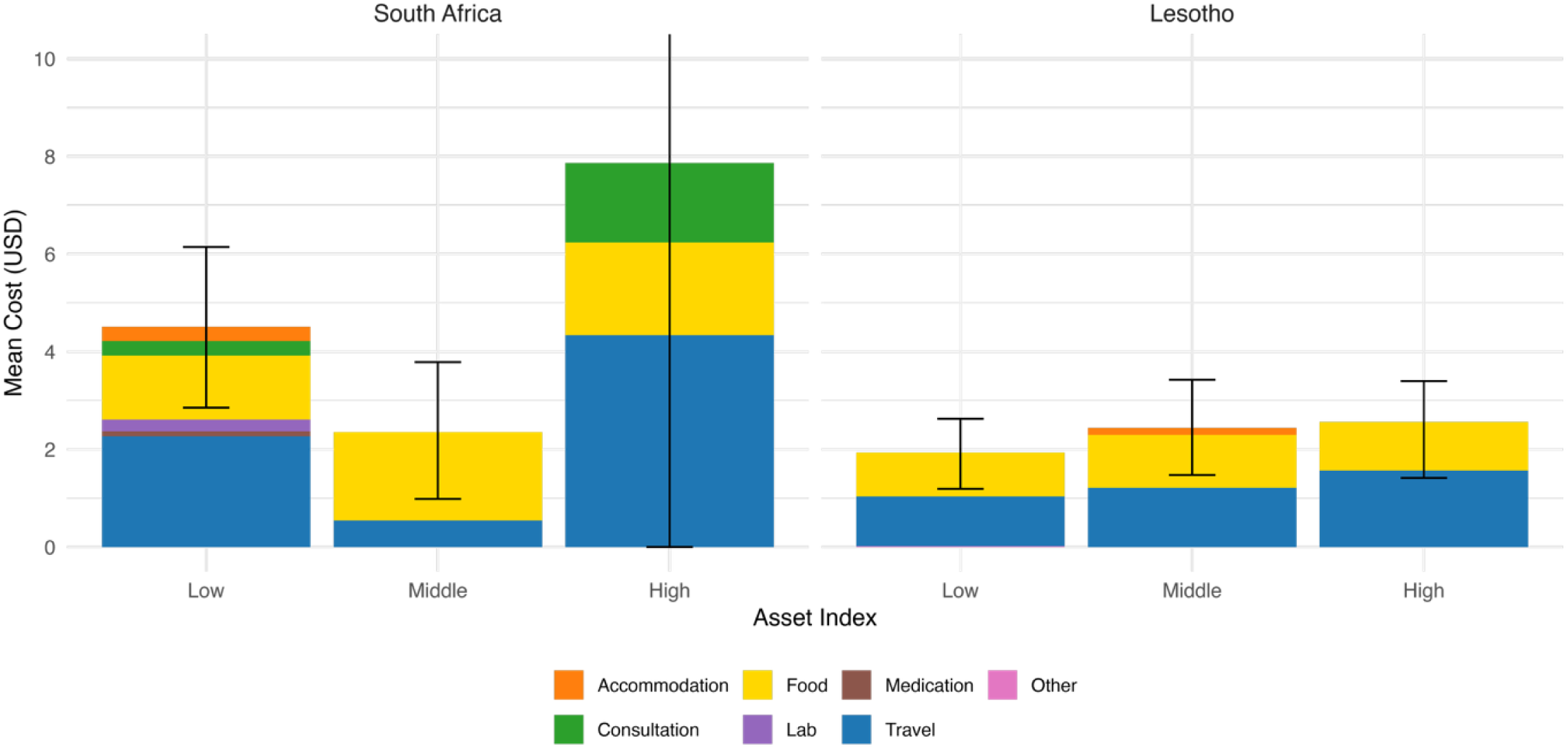
**Average out-of-pocket expenditure to seek TB treatment at follow-up by socioeconomic group and country, disaggregated by expenditure category for TB-positive participants identified via ACF** Note: The out-of-pocket expenditure here refers to the costs incurred by an individual for their most recent TB-related healthcare visit after being diagnosed with TB through active case finding during the follow-up data collection period. The stacked bars illustrate the composition of average expenditure across seven cost categories. Black vertical lines represent 95% confidence intervals around the total mean cost per group. The upper bound for the high SES group in South Africa extends till USD 23.37 and has been clipped for clarity. Costs are collected in ZAR/LSL and converted to USD.

### Household income, welfare support, and perceived financial impact of TB

Baseline monthly household income data were available for 413 participants across the four groups (195 in South Africa and 218 in Lesotho), while follow-up income data were available for 83 TB-positive participants diagnosed via ACF (28 in South Africa and 55 in Lesotho). In total, 29.5% (n = 122) of participants with baseline income data reported receiving social welfare payments, including 98 in South Africa and 24 in Lesotho. In South Africa, the majority of recipients were from the low socioeconomic group (n = 54), followed by the middle (n = 34) and high (n = 10) groups. In Lesotho, 20 recipients were classified as low SES, with only one participant in the middle and three in the high SES groups.

Participants with follow-up income data were similar in socioeconomic profile to those without (Table S8). Panel A of Figure 4 shows the distribution of monthly household income at baseline and follow-up among participants who reported income. Panel B displays the within-household change in income between baseline and follow-up among ACF-identified TB-positive participants. Median monthly household income at baseline was $139.94 (IQR: 86.78 - 253.84) in South Africa and $48.82 (IQR: 27.12 - 108.48) in Lesotho; at follow-up, the corresponding values were $118.24 (IQR: 81.36 - 296.42) and $46.10 (IQR: 0.00 - 54.24), respectively.

**Figure 4.**
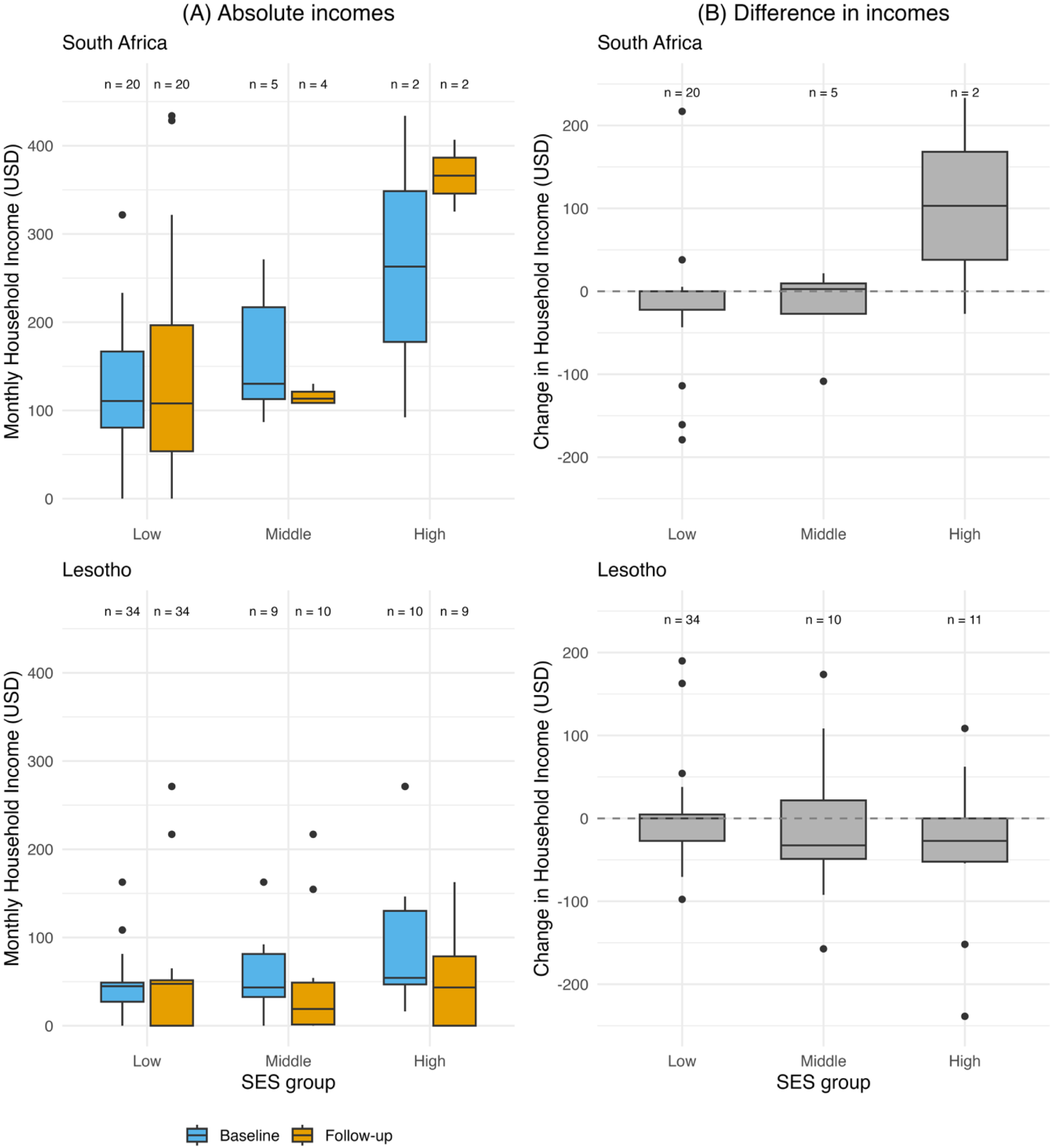
Monthly household income and changes in income by SES group and country. Note: Panel (A) shows absolute monthly household income reported at baseline and follow-up. Panel (B) displays the change in income between baseline and follow-up for the same participants. Observations are stratified by country, socioeconomic group, and time point. All values are presented in USD (1 ZAR = 0.05424 USD). Observations with income > USD 450 were excluded for visual clarity (n = 2 at baseline; n = 3 at follow-up).

No significant differences in baseline household income were found by sex, diagnostic strategy, or TB status (Table S9), though ACF-identified participants had lower absolute income than those diagnosed via PCF, especially in Lesotho (Figure S2). None of the changes in household income were statistically significant across SES groups, countries, or sex (see Supplement). At follow-up, most TB-positive participants with income data (n = 75; 87.2%) reported no perceived change in their household financial situation following the onset of TB symptoms (Table S10). However, 10.47% (n = 9) reported that their household became either poorer or much poorer after their TB diagnosis.

### Economic burden and coping strategies among TB-affected households

To assess financial burden, we calculated the share of monthly income spent on OOP expenses at baseline. Overall, 3 participants (3.3%) incurred OOP costs exceeding 20% of household income (1 in SA, 2 in LS). In South Africa, the median burden increased with SES: 2.7% (IQR: 1.3-6.4%) in the low group, 2.3% (IQR: 1.6-6.1%) in the middle, and 14.6% (IQR: 1.8-20.4%) in the high. In Lesotho, the low SES group had the highest and most variable burden: 3.7% (IQR: 2.5-60.2%). Differences by SES and diagnostic pathway were not statistically significant (see Supplement). Among those reporting OOP expenses, 58.6% in South Africa and 31.0% in Lesotho reported zero OOP spending (Table S11).

Among TB-positive participants with follow-up income data, 6 (7.2%) borrowed or received money for TB-related costs (5 in SA, 1 in LS); 4 were from the low SES group. All support came from family or friends, with a median amount of $2.16 (IQR: $1.60-$2.70).

## Discussion

Our findings highlight important socioeconomic and demographic patterns in TB-associated outcomes, as well as in the financial burden associated with TB-related care, though the strength of these associations warrants cautious interpretation. In South Africa, individuals from wealthier households were more likely to incur higher OOP expenditures and to have documented linkage to care as indicated by the presence of a TB record. This raises the possibility that wealth may facilitate better access to diagnosis and treatment, albeit at a higher financial cost. In contrast, individuals from poorer households in Lesotho appeared to face a higher financial burden relative to income, despite reporting similar or lower absolute expenditures, suggesting a hidden inequity in how costs are experienced and absorbed across income groups.

TB-related outcomes also varied by demographic characteristics: older adults, men, and people living with HIV (PLHIV) had higher odds of TB positivity and treatment uptake, while women were less likely to be TB-positive but significantly more likely to report NCDs. These patterns suggest gendered and age-related dimensions of health-seeking behaviour and disease burden that merit further investigation. Additionally, higher education was positively associated with PCF diagnosis and inversely via ACF, suggesting differences in health literacy, care-seeking autonomy, or even exposure to outreach interventions.

These trends align with previous research documenting elevated TB risk among older individuals and men in South Africa and Lesotho, and across the African region [2, 3, 15–17]. Informal employment has similarly been linked to poorer treatment outcomes, highlighting the vulnerability of socioeconomically disadvantaged groups [6].

HIV prevalence was higher in South Africa than in Lesotho, likely reflecting our study’s implementation in KwaZulu-Natal, the province with the second highest HIV burden nationally [18]. We observed an association between HIV status and the TB diagnostic pathway: 63.7% of individuals identified through PCF were HIV-positive, compared to 24.7% through ACF. This may reflect increased symptom severity among PLHIV and greater care-seeking due to immunosuppression [19]. Additionally, PLHIV are often already engaged in care and are screened more frequently, which may facilitate earlier TB detection and treatment [20]. Hence, the higher odds of PCF identification among PLHIV likely stem from differences in care engagement and symptom burden, rather than the diagnostic strategy itself. Similar findings have been reported in Kenya, where PLHIV were more likely to be diagnosed via PCF due to symptom presence and immunosuppression [21].

Our analysis of OOP expenditures revealed distinct country-level patterns. In South Africa, individuals in the high SES group incurred the highest TB-related costs, driven largely by spending on consultations and travel. This suggests that wealthier households may utilise higher-cost healthcare services, reflecting fewer financial constraints. In Lesotho, by contrast, participants in the low and middle SES group incurred higher absolute OOP expenditures, with hospitalisation and transport as the dominant cost components. Notably, hospitalisation costs were only reported prior to diagnosis and were mostly associated with PCF-based identification, which may indicate more severe illness due to delayed diagnosis. These patterns may reflect differences in health system access points, since most participants in South Africa visited primary clinics while those in Lesotho visited district hospitals. Furthermore, participants from the low SES group may have spent less on food due to limited capacity to pay, rather than lower need. Our findings on OOP expenditure align with previous findings from South Africa, where travel costs averaged $1.69 per TB-related visit [22]. In our study, travel-related expenses were similar at around $1-2 per visit. These results also align with regional estimates of travel costs, which have been found to range from $2-3 per visit [23].

We did not observe any significant change in household income between baseline and follow-up, possibly due to a limited number of participants reporting job loss and an impact on total household earnings. Nearly one-third of participants reported receiving social welfare payments, with uptake concentrated among lower socioeconomic groups in Lesotho. This highlights the role of government support in partially offsetting income vulnerability among TB-affected households – a finding previously documented in literature [24]. Although instances of borrowing and job loss were relatively infrequent, they were disproportionately concentrated among the poorest households, indicating a limited capacity to absorb financial shocks for those in the low SES group. Our findings on monthly household income and the impact of TB-related income losses are consistent with published literature. Previous studies have observed average monthly household income in South Africa to be approximately $103, with income losses due to TB disproportionately affecting lower-income households [25]. Prior studies have shown that the relative burden of TB-related OOP spending is highest among the poorest households in sub-Saharan Africa [26].

Catastrophic healthcare expenditure (CHE) is typically defined as total TB-related expenditures exceeding 20% of a household’s annual income [27, 28]. Although our study assessed only initial TB-related OOP costs relative to monthly income, we observed that just 3.3% of participants incurred expenses above this threshold. Several factors may help explain the low frequency of catastrophic health expenditure in our sample. First, we captured costs from only a single visit prior to diagnosis and from a relatively short follow-up period, potentially underestimating subsequent treatment-related costs. Second, the majority of individuals accessed care at public sector facilities where services are heavily subsidised or free at point of care, limiting direct OOP spending. Lastly, participants diagnosed via ACF may have received more decentralised or community-based services, reducing travel and consultation-related costs. Collectively, these factors may have buffered households from experiencing high initial TB-related financial burden, though longer-term monitoring is needed to fully assess economic impact over the course of illness and treatment. It is important to note that low initial financial burden does not rule out the possibility of future economic vulnerability, particularly in cases of recurrent or drug-resistant TB, where prolonged treatment and income loss can lead to substantial accumulated costs and financial hardship [29].

### Strengths and limitations

A key strength of this study is the reliance on data from a pragmatic, community-based TB diagnostic trial in two high-burden settings. By including TB-positive participants identified through both ACF and PCF strategies, alongside those without TB, the study allows for a detailed comparison for assessing how socioeconomic status influences diagnostic pathways and access to care. Simultaneously capturing OOP expenditures, household income, and coping strategies, it provides a comprehensive assessment of the early economic burden of TB. The use of a harmonised, national-based IWI to define SES groups strengthens comparability between our study sample and the general populations of South Africa and Lesotho.

To our knowledge, this is one of few empirical studies to measure income loss among TB patients during the early stages of treatment. Most previous research has either assessed costs across the full treatment duration or relied on modelled estimates [28]. Other studies have found that income losses peak at treatment completion but stagnate thereafter - highlighting the importance of sustained research and support to mitigate TB’s long-term impact on household poverty [30].

Despite its strengths, this study has some limitations. First, the overall sample size was modest, particularly when stratified by SES, country, or diagnostic pathway, limiting statistical power for subgroup analyses. Second, follow-up income data were only available for ACF-identified TB-positive participants, reducing comparability across diagnostic strategies. Third, the 56-day follow-up period may have been too short to detect longer-term economic consequences of TB. Fourth, reliance on self-reported data for income, OOP, and comorbidities are subject to potential recall and reporting biases. Fifth, our estimates of financial burden are based on monthly income and are therefore not directly comparable to standard definitions of CHE, which typically rely on annual income and total treatment costs [27]. Finally, educational attainment may have been misreported due to inconsistent interpretations of schooling years across settings. For example, while 73% of our South African sample reported 7-12 years of schooling, other studies report that only 57% have completed primary education [15].

## Conclusion

Our study results indicate the need for more targeted and equitable strategies to reduce the health and economic burden of TB. The higher TB positivity and lower treatment uptake observed among older adults and men suggest that these groups should be prioritised in active case-finding and outreach strategies, particularly in settings where they may underutilise health services. At the same time, the disproportionately high NCD burden among women warrants greater attention to the TB-NCD comorbidity, which has been shown to result in worse health outcomes than TB alone [31]. This highlights the importance of integrating NCD screening and management into TB care platforms, especially for women. Country-level differences in healthcare access - with most participants in South Africa visiting primary healthcare clinics and those in Lesotho attending hospitals - point to the need to strengthen decentralised, community-based care in Lesotho and to improve referral systems in South Africa to promote more equitable service delivery. Finally, although only a small proportion of participants experienced catastrophic costs, income loss and borrowing were disproportionately concentrated among the poorest households. This underscores the importance of financial protection mechanisms such as transport stipends or targeted welfare support to cushion the economic impact of TB. Future research should assess the longer-term financial consequences of TB illness across socioeconomic groups and evaluate the effectiveness of integrated care and social protection interventions in improving both health and economic outcomes.

## Data Availability

The de-identified dataset supporting the conclusions of this study will be deposited in Zenodo upon acceptance for publication. All identifying information has been removed, and ethical approvals allow for data sharing under these conditions. A provisional DOI and private reviewer link will be provided during peer review.

## Acknowledgements

We are sincerely grateful to the field workers and all participants in the TB TRIAGE+ trial whose invaluable time, trust, and experiences made this research possible.

## Notes

### Competing Interest Statement

The authors have declared no competing interest.

### Clinical Trial

ClinicalTrials.gov (NCT05526885), South African National Clinical Trials Register (SANCTR DOH-27-092022-8096).

### Clinical Protocols

https://bmjopen.bmj.com/content/15/7/e093989

### Author Declarations

The study protocol was approved by the National Health Research Ethics Committee in Lesotho (NH-REC, ID52-2022), the Human Sciences Research Council Research Ethics Committee in South Africa (HSRC REC, REC 2/23/09/20), the Provincial Health Research Committee of the Department of Health of KwaZulu-Natal (KZ_202209_022), and the Swiss Ethics Committee Northwest and Central Switzerland (EKNZ, AO_2022–00044). Written informed consent was obtained from all study participants prior to enrolment.

